# Alzheimer’s risk markers and resting-state dynamic functional connectivity: Cross-Sectional Findings from the AGUEDA Study

**DOI:** 10.64898/2026.02.24.26346860

**Authors:** Andrea Coca-Pulido, Patricio Solis-Urra, Oren Contreras-Rodríguez, Carles Biarnés, Marcos Olvera-Rojas, Shivangi Jain, Anuradha Sehrawat, Yihun Chen, Yolanda García-Rivero, Manuel Gomez-Rio, Kirk I. Erickson, Jose Mora-Gonzalez, Irene Esteban-Cornejo

## Abstract

**Background and Objectives:** Alzheimer’s disease (AD) is characterized by early disruptions in brain connectivity. However, how genetic and biological markers of AD risk relate to dynamic functional connectivity (dFC) remains unclear. This study examined whether AD-related pathology, genetic risk, and blood-based biomarkers (BBMs) of neurodegeneration are associated with local and distant resting-state dFC patterns, and whether these relate to cognitive performance in cognitively normal older adults.

**Research Design and Methods:** We analyzed baseline data from 86 cognitively normal older adults (71.6 ± 3.9 years; 60.5% female) enrolled in the AGUEDA trial (NCT05186090). Participants underwent Aβ-PET imaging, APOE4 genotyping, and plasma quantification of BBMs (Aβ42/40, BD-tau, GFAP, NfL, p-tau181, p-tau217). Resting-state fMRI was used to compute voxel-wise local and distant dFC using a stepwise connectivity framework. General linear models tested associations between AD pathology, APOE4 status, and BBMs with dFC, adjusting for age, sex, and education. Additional models examined links between dFC and six cognitive domains

**Results:** Aβ-positive individuals and APOE4 carriers showed lower local connectivity in frontal regions, while APOE4 carriers exhibited higher distant connectivity in the superior motor area, inferior frontal gyrus, and anterior insula. Among BBMs, only neurofilament light chain (NfL) was associated with both lower local (insula, cingulate) and higher distant (precuneus, putamen, thalamus, supramarginal, superior motor area) connectivity. Regions showing higher distant connectivity related to APOE4 or NfL were associated with poorer cognitive performance.

**Discussion and Implications:** Dynamic functional connectivity reveals early network alterations in AD risk, characterized by reduced local and elevated distant connectivity—patterns linked to poorer cognition and potential early neurofunctional vulnerability in aging.

## 1. Introduction

As life expectancy increases, dementia prevalence continues to rise, posing a major public health challenge ^1^. Alzheimer’s disease, the leading cause of dementia, is characterized by amyloid-beta plaques and neurofibrillary tangles of hyperphosphorylated tau, which promote synaptic dysfunction, neuronal loss, and cognitive decline ^2,3^. Although positron emission tomography enables in vivo detection of these pathologies, its clinical use is limited by cost and invasiveness. More accessible tools have emerged in the form of blood-based biomarkers, which provide reliable indices of Alzheimer’s disease pathology. Plasma amyloid-beta 42/40 ratio and phosphorylated tau 217 show strong diagnostic accuracy even in early stages ^4^, while neurofilament light chain, glial fibrillary acidic protein, and brain-derived tau capture broader neurodegenerative processes, reflecting axonal injury and astrocytic reactivity ^5^. In parallel, carrying the apolipoprotein E ε4 allele confers a three- to four-fold increased risk of late-onset Alzheimer’s disease ^6^.

Beyond biological markers, functional magnetic resonance imaging provides insights into how underlying pathology affects brain network organization. Resting-state functional connectivity reflects synchronized activity between brain regions and serves as a sensitive marker of neural communication ^7,8^. In healthy aging, networks maintain a modular architecture that balances local segregation, supporting specialized processing, with distant integration, enabling efficient communication across distributed regions ^9^. In Alzheimer’s disease, this balance is disrupted: patients show reduced local segregation and increased distant integration, resulting in fragmented and inefficient networks ^10,11^, alterations that are closely linked to impaired cognition ^12^. Similar, though less severe, patterns are observed in aging, where reduced within-network connectivity and increased between-network connectivity have been associated with poorer memory, executive function, and diminished network specialization ^13–15^. Despite advances, methodological gaps remain. Most prior studies relied on predefined regions, seed-based methods, or static connectivity analyses, which limit the capacity to capture whole-brain and dynamic changes ^16–18^. Whole-brain approaches allow a more comprehensive and unbiased characterization of connectivity ^19^, and considering connectivity as dynamic rather than static may offer a more sensitive window into early alterations ^20,21^. Yet, no previous studies have examined how plasma biomarkers of Alzheimer’s disease pathology and neurodegeneration relate to dynamic functional connectivity.

To address this gap, we employed a whole-brain dynamic functional connectivity approach in cognitively normal older adults. We investigated whether amyloid status, apolipoprotein E ε4 carriage, and blood-based markers of neurodegeneration (amyloid-beta 42/40 ratio, brain-derived tau, glial fibrillary acidic protein, neurofilament light chain, phosphorylated tau 181, and phosphorylated tau 217) are associated with local and distant connectivity patterns. We further examined whether these connectivity patterns are linked to cognitive performance across multiple domains. We hypothesized that individuals at greater Alzheimer’s disease risk would show reduced local and/or increased distant connectivity, and that such alterations would be associated with poorer cognition.

## 2. Research Design and Methods

### 2.1. Study design and participants

A total of 91 cognitively normal older adults were recruited for the “Active Gains in brain Using Exercise During Aging” (AGUEDA) trial (ClinicalTrials.gov: <u>(NCT05186090)</u>), a randomized controlled trial evaluating a 24-week resistance exercise program on executive function ^22^. Participants were recruited from Granada (Spain). Full inclusion/exclusion criteria are reported elsewhere ^23^. Briefly, eligibility required: (i) age 65–80 years; (ii) physical inactivity (no resistance training in past six months or <600 MET-min/week on the International Physical Activity Questionnaire); (iii) no cognitive impairment (STICS-m ≥ 26, MMSE ≥ 25, MoCA ≥ 24 for <71 years, ≥ 22 for 71–75 years, ≥ 21 for >75 years); and (iv) no relevant depressive symptoms (GDS <15). All instruments were administered in validated Spanish versions. For this cross-sectional analysis, we used baseline data collected between March 2021 and May 2022. Five participants were excluded due to excessive motion or invalid slices, leaving 86 for analysis.

The study followed the Declaration of Helsinki and was approved by the Ethics Board of the Andalusian Health Service (CEIM/CEI Provincial de Granada; #2317-N-19). Written informed consent was obtained from all participants.

### 2.2. AD Pathological Status: Brain Aβ Deposition

Aβ deposition was assessed with a Biograph-Vision 600 Edge PET/CT scanner (Siemens, Erlangen, Germany) at the Hospital Universitario Virgen de las Nieves (HUVN) (Granada, Spain). The tracer [18F] Florbetaben (Neuraceq; Piramal Pharma) was administered intravenously. Images were acquired 90–110 min post-injection of 300 MBq ± 20% ^24^ following a standardized acquisition and processing protocol ^25–27^. Additional PET parameters are reported elsewhere ^23^. Participants were classified as Aβ-negative (CL <12) or Aβ-positive (CL ≥12). Details on BBMs processing are reported elsewhere (Olvera-Rojas et al., submitted).

### 2.3. AD Genetic Risk: APOE4 Status

Blood samples were collected under fasting conditions (08:00–10:00 a.m.) at the HUVN in Granada, Spain. APOE4 status was determined by detecting at least one ε4 allele from two single-nucleotide polymorphisms (rs7412, rs429358) using TaqMan assays. Participants were classified as carriers or non-carriers. Further genotyping details are available elsewhere (Olvera-Rojas et al., submitted).

### 2.4. BBMs of neurodegeneration

Fasting blood samples were collected following the same protocol described in Section 2.3. Plasma levels of BBMs (ie. Aβ40/Aβ42 ratio, BD-tau, GFAP, NfL, p-tau181, and p-tau217) were used as indicators of neurodegeneration. The Aβ42/Aβ40 ratio was determined using both Single Molecule Array (SIMOA) and Immunoprecipitation Mass Spectrometry (IPMS) technologies, and was calculated as Aβ42 divided by Aβ40 and multiplied by –1, such that higher values reflected greater amyloid burden. Further details on BBMs of neurodegeneration processing are available elsewhere (Olvera-Rojas et al., submitted).

### 2.5. Magnetic Resonance Imaging (MRI) procedure

#### 2.5.1. Data acquisition

A 3.0 T Siemens Magnetom Prisma Fit scanner (Siemens Medical Solutions, Erlangen, Germany) with a 64-channel head coil was used for image acquisition. Rs echo-planar image (EPI) sequence was used to collect functional images. Participants were instructed to fix the gaze at a white cross in black background during acquisition. The acquisition parameters for the rs EPI sequence were as follow: repetition time (TR) = 1000 ms, echo time (TE) = 40 ms, multiband factor = 8 (center for magnetic resonance research EPI sequence), resolution = 2.5 × 2.5 × 2.5 mm, 64 slices, and a scan duration of 8 min and 12 s. T1-weighted structural images, used for normalization, were acquired with the following parameters: TR = 2400 ms, TE = 2.31 ms, inversion time = 1060 ms, field of view = 256 mm, voxel size = 0.8 × 0.8 × 0.8 mm, 224 slices, and scan duration = 6 min and 38 s. Further information regarding image acquisition protocols can be found elsewhere ^23^.

#### 2.5.2. Functional image processing

##### 2.5.2.1. First level analysis - Preprocessing of rs EPI images

Functional imaging data were preprocessed using FSL 6.0.7.4, Matlab 2022a, and the CONN 22a toolbox in the Neurodesk platform; an accessible, flexible and portable data analysis environment for reproducible neuroimaging.

###### Creation of fieldmaps

The FSL software was implemented for the creation of fieldmaps by combining the anterior-posterior and posterior-anterior EPI images. These fieldmaps were used to correct susceptibility-induced distortions in fMRI, which arise due to magnetic field inhomogeneities, particularly in regions near air-tissue interfaces such as the sinuses or the base of the skull. This method, known as *topup*, estimates a field distortion map that can be applied to correct the geometry of distorted images, improving the accuracy of subsequent analyses.

###### Smoothing and denoising

The CONN 22a toolbox was implemented for preprocessing the rs EPI images. The pipeline used was the *“Preprocessing pipeline for volume-based analyses (indirect normalization to MNI-space) when fieldmaps are available”.* This pipeline accomplish the following steps: (1) label current functional files as “original data”, (2) creation of voxel-displacement maps for susceptibility distortion correction, (3) realignment with susceptibility distortion correction using fieldmaps previously uploaded to the software (subject motion estimation and correction), (4) center functional images to (0,0,0) coordinates (translation), (5) outlier detection (ART-based identification of outlier scans for scrubbing), (6) indirect segmentation and MNI-space normalization (simultaneous Gray/White/CSF segmentation and MNI normalization), (7) label current functional files as “mni-space data”, (8) center structural images to (0,0,0) coordinates (translation), (9) structural segmentation and normalization (simultaneous Gray/White/CSF segmentation and MNI normalization), (10) smoothing (spatial convolution with Gaussian kernel of 6 mm), and (11) label current functional files as “smoothed data”. The step where slice timing correction is used for interslice differences in acquisition time was skipped as the TR was short (TR=1s) and it was a multi-band acquisition. This means that the temporal differences between slices are minimal, making slice timing correction step unnecessary. This preprocessing pipeline resulted in smoothed rs EPI images.

The smoothed data was denoised by setting a band-pass filter of 0.008–0.09 Hz allowing the retention of meaningful rs BOLD signal fluctuations while minimizing noise from non-neuronal sources. The band-pass filter selection was adjusted based on the window size used for functional connectivity analysis, which in turn depends on the TR of the acquisition. The lower bound (0.008 Hz) was determined as the inverse of the selected window size, ensuring that only relevant low-frequency fluctuations are preserved. The upper bound (0.09 Hz) was maintained to eliminate high-frequency noise associated with physiological and scanner-related artifacts ^28^.

###### Reslicing and resampling

Reslicing and resampling were performed using Matlab 2022a ^29^ to ensure alignment of functional images with a standard reference template (*avg152T1_gray_bin* nii file). This process standardized voxel resolution (from 2.4×2.4×2.4mm to 6.0×6.0×6.0mm) and spatial orientation across subjects, facilitating accurate group-level analyses and rs dFC computations.

###### Quality control

As part of the quality control process, several indicators were used to identify potential outliers. These included the percentage of invalid volumes, maximum motion (cutoff >□6□mm), mean motion (cutoff >□1.5□mm), and framewise displacement (cutoff >□1.5□mm) extracted from each preprocessed functional image. We also conducted a visual inspection of violin and histogram plots from the Quality Assurance section, displaying the distribution of motion and global signal change values across subjects. Additionally, potential outliers were flagged using the 1.5□×□interquartile range rule beyond the third quartile. Among all these criteria, the percentage of invalid slices was considered the primary exclusion criterion. Five participants with more than 25% invalid volumes were excluded from further analyses to ensure sufficient data quality, in accordance with established control standards for rs data preprocessing ^30^. Based on this process, five participants were classified as outliers and removed from subsequent analyses.

###### Rs dFC maps computation

Rs dFC maps were computed using Matlab 2022a ^29^ and a pipeline based on sliding window functional brain connectivity analysis. For each subject, voxel-wise time series were extracted from denoised data. Rs dFC was estimated using a 20s sliding window, which moved 7s forward at each step along the time series, allowing overlapping segments for capturing dynamic changes in connectivity ^28^. Pearson correlation matrices were computed for each window and transformed using Fisher’s z-transformation to enhance statistical normality. Negative correlations and non-significant values (p > 0.05) were removed to retain meaningful connectivity patterns.

To refine the rs dFC structure, a stepwise functional brain connectivity analysis, as described by Sepulcre et al. (2018), was implemented ^28^. This approach iteratively expands connectivity patterns by progressively multiplying the rs dFC graph, preserving only recurrent and stable network interactions while filtering out transient or spurious connections. The stepwise functional brain connectivity model enables the identification of hierarchical connectivity pathways, distinguishing, local connectivity, which captures short-range, recurrent, and stepwise interactions within functional modules, and distant connectivity, which represents long-range integrative connections between voxels. The resulting local and distant connectivity maps were used as primary neuroimaging outcomes in subsequent group comparisons and linear regression analyses.

### 2.6. Cognitive Function

Cognitive function was assessed using six domain-specific composite scores: executive function, episodic memory, processing speed, visuospatial processing, working memory, and attentional/inhibitory control. The executive function composite was derived using confirmatory factor analysis conducted in this sample, while the additional five cognitive domains were computed based on the multi-factor confirmatory factor analysis model developed in the IGNITE study, a RCT with a considerably larger sample size, similar objectives, participants’ characteristics, and cognitive tests ^31^. In all domains, higher scores indicate better cognitive performance. Further details on cognitive domains creation are available elsewhere (Fernández-Gamez et al., in review).

### 2.7. Statistical Analysis

The characteristics of the study sample are presented as means and standard deviations (SD). To assess normality, a visual inspection of histograms, p-values (p ≤ 0.05) from the Kolmogorov–Smirnov test, and Q–Q (quantile–quantile) plots were reviewed. Based on these assessments, most variables did not require transformation; however, p-tau217, p-tau181, GFAP, and NfL underwent logarithmic transformation to achieve a normal distribution. Missing data were present for a small subset of participants across different variables. Specifically, one participant had missing data for BD-tau, one participant for p-tau217, one participant for Aβ40/42 (SIMOA), one participant for both GFAP and NfL, and one participant for executive function. Due to the low proportion of missing data, no imputation was performed ^32^. Therefore, analyses involving these variables were performed with the corresponding number of participants with valid data.

First, we conducted general linear models in SPM to examine group differences in local and distant rs dFC patterns across AD pathology (i.e., Aβ-positive vs. Aβ-negative) and genetic status (i.e., APOE4 carriers vs. non-carriers). Age, sex assigned at birth and years of education were included in analyses as covariates based on previous literature ^33–35^. c

Second, we employed individual linear regression models to examine the association between BBMs of neurodegeneration (i.e., Aβ40/42 (SIMOA), Aβ40/42 (IPMS), BD-tau, GFAP, NfL, p-tau181, p-tau217) and rs dFC, while controlling for age, sex, and education. Both analyses were conducted using Statistical Parametric Mapping software (SPM12; Wellcome Department of Cognitive Neurology, MA). Statistical significance was set at a peak-level uncorrected p-value (p ≤ 0.01) and a cluster-level FWE-corrected p-value (pFWE < 0.05) with corrections applied within a gray matter mask derived from SPM12 tissue probability maps to assess the relationship between predictor variables and connectivity maps. Then, we extracted the attractor strength from the peak coordinates of each significant cluster using SPM12 and calculated the standardized β values using RStudio for Mac (Version 2023.03.1, lm.beta).

Lastly, a third set of linear regressions was performed to determine whether brain regions showing significant group differences for rs dFC, based on Aβ pathology status (i.e., Aβ-positive vs. Aβ-negative) and genetic status (i.e., ApoE ε4 carriers vs. non-carriers), or associations with BBMs of neurodegeneration were also linked to cognitive function (i.e., executive function, episodic memory, processing speed, visuospatial processing, working memory, attentional/inhibitory control). These analyses were conducted using RStudio for Mac (Version 2023.03.1 + 446)

## 3. Results

### 3.1. Descriptive Characteristics

The descriptive characteristics of the study sample are shown in Table 1. The participants had a mean age of 71.63 ± 3.94 years. The sample consisted of 60.47% females. Regarding AD pathology, 19 participants tested positive for Aβ (CL>12), and in terms of AD genetic status, 11 participants were APOE4 carriers.

**Table 1.**
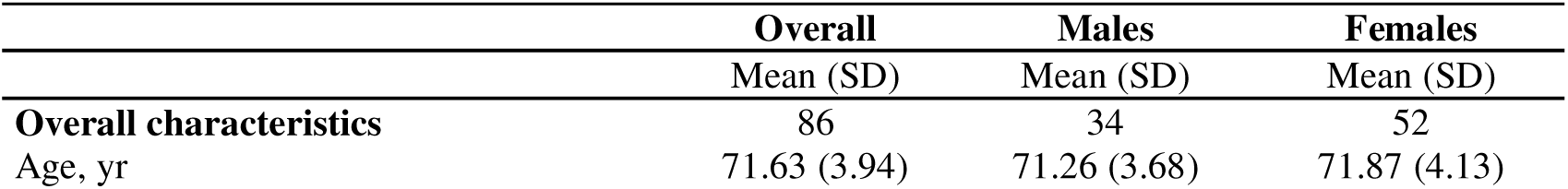
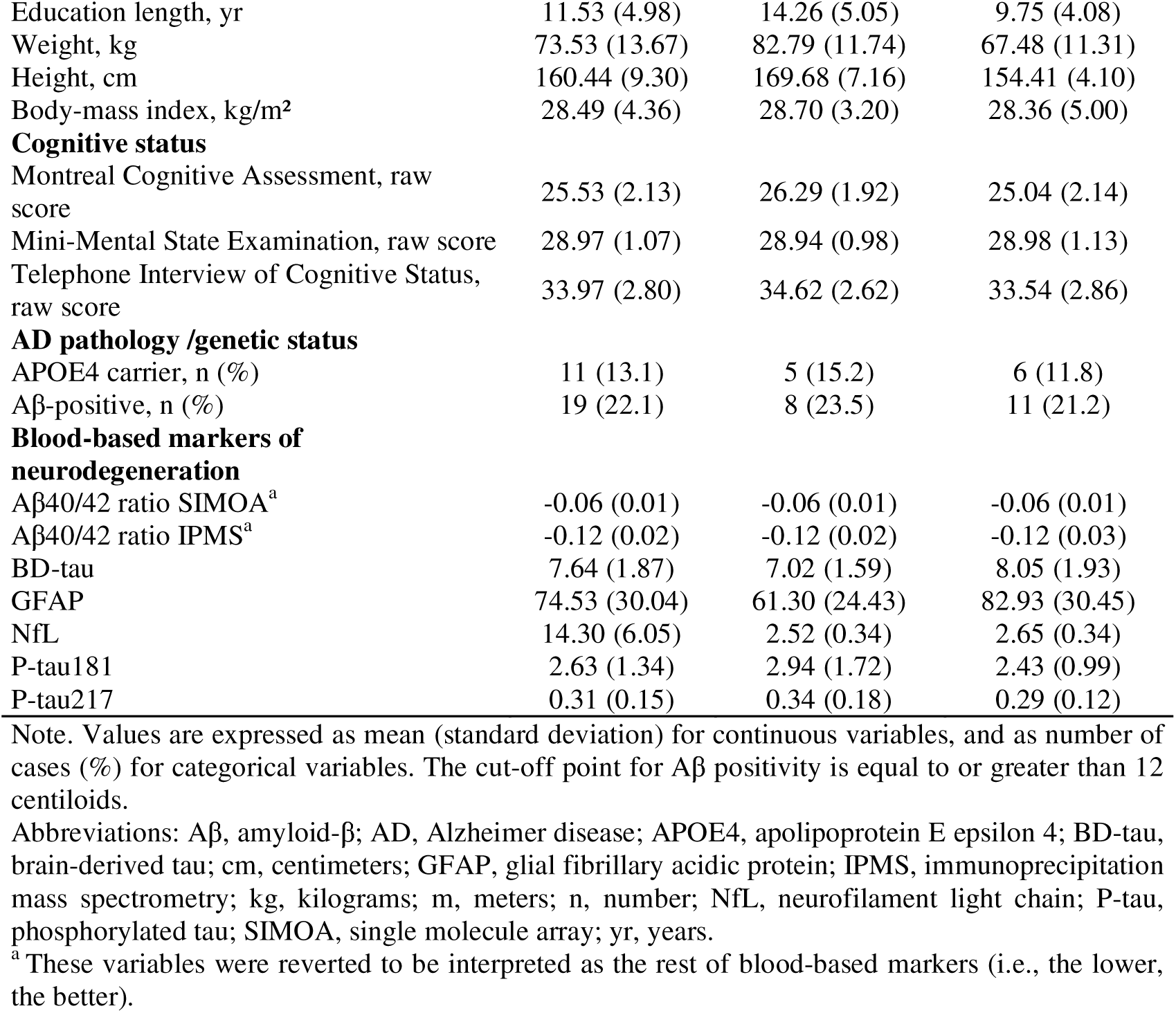
Descriptive characteristics of the study sample (n=86)

|  | **Overall** | **Males** | **Females** |
| --- | --- | --- | --- |
|  | Mean (SD) | Mean (SD) | Mean (SD) |
| **Overall characteristics** | 86 | 34 | 52 |
| Age, yr | 71.63 (3.94) | 71.26 (3.68) | 71.87 (4.13) |
| Education length, yr | 11.53 (4.98) | 14.26 (5.05) | 9.75 (4.08) |
| Weight, kg | 73.53 (13.67) | 82.79 (11.74) | 67.48 (11.31) |
| Height, cm | 160.44 (9.30) | 169.68 (7.16) | 154.41 (4.10) |
| Body-mass index, kg/m<sup>2</sup> | 28.49 (4.36) | 28.70 (3.20) | 28.36 (5.00) |
| **Cognitive status** |  |  |  |
| Montreal Cognitive Assessment, raw score | 25.53 (2.13) | 26.29 (1.92) | 25.04 (2.14) |
| Mini-Mental State Examination, raw score | 28.97 (1.07) | 28.94 (0.98) | 28.98 (1.13) |
| Telephone Interview of Cognitive Status, raw score | 33.97 (2.80) | 34.62 (2.62) | 33.54 (2.86) |
| **AD pathology /genetic status** |  |  |  |
| APOE4 carrier, n (%) | 11 (13.1) | 5 (15.2) | 6 (11.8) |
| Aβ-positive, n (%) | 19 (22.1) | 8 (23.5) | 11 (21.2) |
| **Blood-based markers of neurodegeneration** |  |  |  |
| Aβ40/42 ratio SIMOA<sup>a</sup> | -0.06 (0.01) | -0.06 (0.01) | -0.06 (0.01) |
| Aβ40/42 ratio IPMS<sup>a</sup> | -0.12 (0.02) | -0.12 (0.02) | -0.12 (0.03) |
| BD-tau | 7.64 (1.87) | 7.02 (1.59) | 8.05 (1.93) |
| GFAP | 74.53 (30.04) | 61.30 (24.43) | 82.93 (30.45) |
| NfL | 14.30 (6.05) | 2.52 (0.34) | 2.65 (0.34) |
| P-tau181 | 2.63 (1.34) | 2.94 (1.72) | 2.43 (0.99) |
| P-tau217 | 0.31 (0.15) | 0.34 (0.18) | 0.29 (0.12) |

Note. Values are expressed as mean (standard deviation) for continuous variables, and as number of cases (%) for categorical variables. The cut-off point for Aβ positivity is equal to or greater than 12 centiloids.
Abbreviations: Aβ , amyloid-β ; AD, Alzheimer disease; APOE4, apolipoprotein E epsilon 4; BD-tau, brain-derived tau; cm, centimeters; GFAP, glial fibrillary acidic protein; IPMS, immunoprecipitation mass spectrometry; kg, kilograms; m, meters; n, number; NfL, neurofilament light chain; P-tau, phosphorylated tau; SIMOA, single molecule array; yr, years.
<sup>a</sup>These variables were reverted to be interpreted as the rest of blood-based markers (i.e., the lower, the better).

### 3.2. Differences in rs dFC among AD pathology / genetic status

Figure 1 (and Table S1 for numeric data) presents cluster located in brain regions showing significant group differences in local (A) and distant (B) rs dFC patterns based on AD pathology (Aβ status) and genetic status (APOE4), controlling for covariates. Regarding local connectivity, APOE4 carriers showed significant lower local connectivity in a cluster located in the left inferior frontal gyrus (β = −0.215, k = 40, p = 0.003) compared to APOE4 non-carriers. Similarly, Aβ-positive participants showed significant lower local connectivity in a cluster located in the left inferior frontal gyrus (β = −0.047, k = 56, p = 0.001) compared to Aβ negative participants. Regarding distant connectivity, APOE4 carriers showed higher distant connectivity in clusters located in the left Superior Motor Area (β = 0.577, k = 73, p <0.001), left Inferior Frontal Gyrus (β = 0.65, k = 102, p = 0.000), and right Anterior Insula (β = 0.682, k=41, p = 0.011) compared to APOE4 non-carriers. No significant differences were found on AD pathology status for distant connectivity.

**Figure 1.**
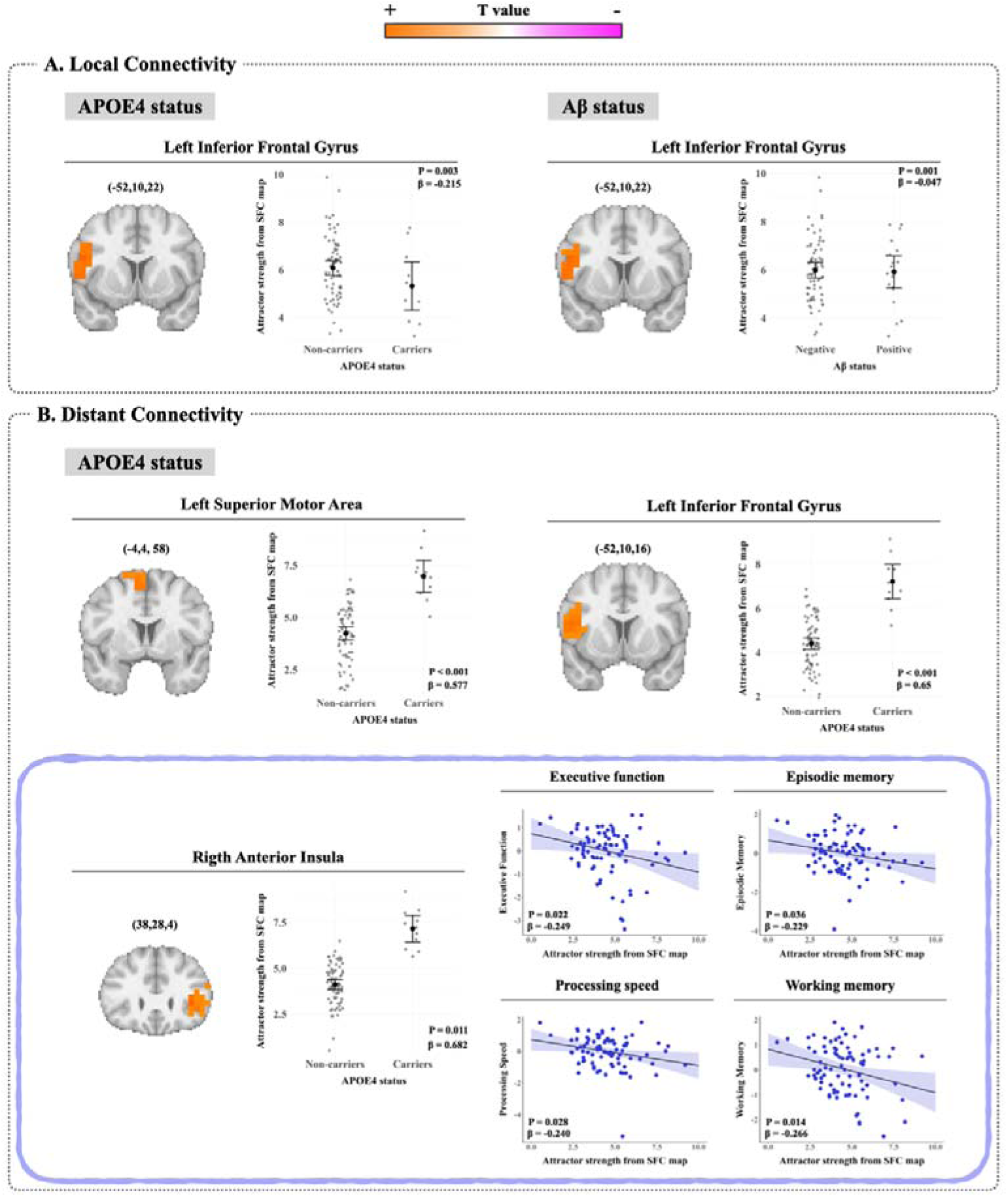
Local and distant brain connectivity patterns associated with AD pathology, genetic status, and cognition. Clusters located in brain regions showing stable local (A) and distant (B) connectivity patterns (attractors) through resting state (rs) dynamic functional brain connectivity (dFC) when associated to Alzheimer Disease pathology/genetic status (predictors) and their further association with cognitive function in cognitively normal older adults (n = 86). Results from graph theory-based rs dFC STEWS method. Age, sex, and education were included as covariates (see Supplementary Table 1). Group scatter plots depict the differences in rs dFC maps among AD pathology and genetic status. Five participants were defined as outliers and excluded from the analyses due to having more than 25% of invalid volumes in the fMRI sequence. Statistical significance was set at a peak-level p-value (uncorrected) ≤ 0.01 and a pFWE(corrected) cluster level < 0.05 to assess differences between groups. Analyses were restricted to voxels within a gray matter mask derived from SPM12 tissue probability maps. The color bar represents T values, which reflect the magnitude and direction of group differences, with lighter orange color indicating regions where the first group in the contrast shows higher values (positive T), and lighter fuchsia color indicating regions where the first group shows lower values (negative T). Anatomical coordinates (X, Y, Z) are given in Montreal Neurological Institute (MNI) Atlas space. P values represent the pFWE(corrected) - cluster level of linear regressions. Standardized beta (β) values quantify the strength and direction of the association. Scatter plots represent individual raw values of attractor strength derived from stepwise functional connectivity (SFC) maps for each participant within the identified clusters. Large black dots indicate group means, and error bars represent 95% confidence intervals of the mean. Gray dots represent individual data points (raw values). The additional blue scatter plots represent clusters showing further associations with cognitive function. No other associations between attractor strength and cognitive function were found (see Supplementary Table 3). APOE4 allele status: carriers vs non-carriers. Aβ status: positive for Amyloid-β vs negative. Attractor strength refers to the weighted convergence of functional connectivity streams across multiple-step paths, computed separately for local and distant connectivity as described in the Methods section.

### 3.3. Association of BBMs of neurodegeneration with stable local and distant rs dFC

Figure 2 (and Table S2 for numeric data) presents significant associations between BBMs of neurodegeneration and clusters located in brain regions with stable local (Figure 2A) and distant (2B) rs dFC patterns (attractors) controlling for covariates. Regarding local connectivity, participants with higher levels of NfL showed significant lower local connectivity in clusters located in the right Middle Cingulate (β = −0.414, k=28, p =0.027) and right Insula (β = −0.453, k=38, p =0.005). Regarding distant connectivity, participants with higher levels of NfL showed significant higher distant connectivity in clusters located in the right Superior Motor Area (β = 0.389, k=98, p < 0.001), right Putamen (β = 0.439, k=181, p < 0.001), right Thalamus (β = 0.423, k=30, p = 0.048), left Supramarginal (β = 0.402, k=119, p < 0.001), and right Precuneus (β = 0.430, k=40, p = 0.012). No significant associations were found for Aβ40/42 ratio (SIMOA), Aβ40/42 ratio (IPMS), p-tau181 and p-tau217 in relation with local or distant connectivity (p > 0.05).

**Figure 2.**
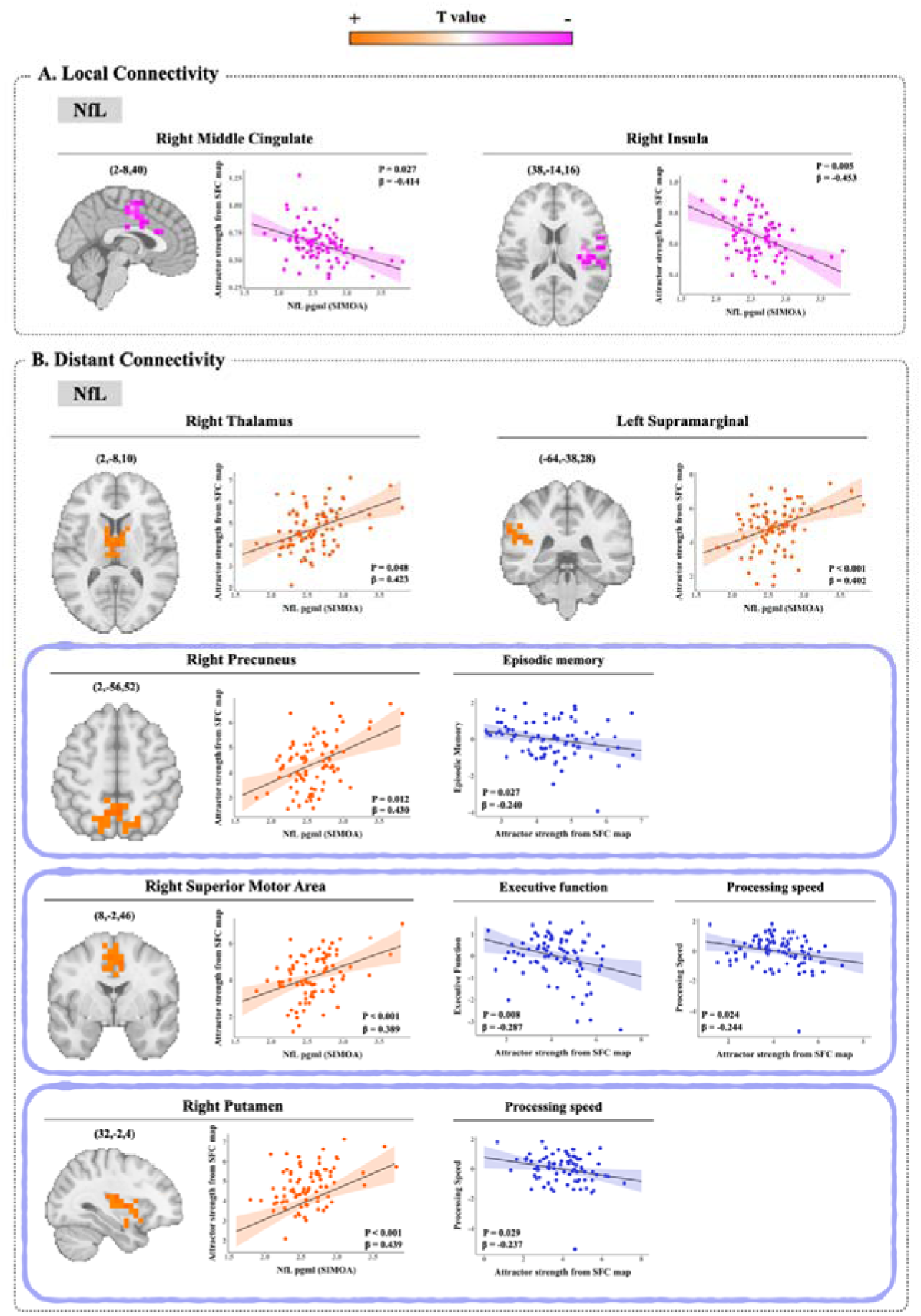
Brain connectivity patterns associated with NfL levels and cognitive function. Clusters located in brain regions showing stable local (A) and distant (B) connectivity patterns (attractors) through resting-state (rs) dynamic functional connectivity (dFC) when associated to NfL (predictors); and their further association with cognitive function in cognitively normal older adults (n = 86). Results from graph theory-based rs dFC STEWS method. Age, sex, and education were included as covariates (see Supplementary Table 2). Five participants were defined as outliers and excluded from the analyses due to having more than 25% of invalid volumes in the fMRI sequence. Statistical significance was set at a peak-level p-value (uncorrected) ≤ 0.01 and a pFWE(corrected) cluster level <0.05 to assess associations. Analyses were restricted to voxels within a gray matter mask derived from SPM12 tissue probability maps. The color bar represents T values, with lighter orange color indicating higher significant positive (+) association and lighter fucsia color indicating higher significant negative (−) association Anatomical coordinates (X, Y, Z) are given in Montreal Neurological Institute (MNI) Atlas space. P values represent the pFWE(corrected) - cluster level of linear regressions. Standardized beta (β) values quantify the strength and direction of the association. Scatter plots display individual raw values of attractor strength derived from stepwise functional connectivity (SFC) maps and either NfL levels (pg/ml) or cognitive test scores. Regression lines indicate the direction of the association, and shaded areas represent 95% confidence intervals. Linear regression models were used to illustrate associations between NfL levels and attractor strength, as well as between attractor strength and cognitive function. Attractor strength refers to the weighted convergence of functional connectivity streams across multiple-step paths, computed separately for local and distant connectivity as described in the Methods section. No other associations between attractor strength and cognitive function were found (see Supplementary Table 3). NfL: Neurofilament Light chain.

### 3.4. AD risk markers-related connectivity and their association with cognitive function

Clusters located in brain regions showing significant associations between AD pathology and genetic status-related connectivity and cognitive function are presented in Figure 1B (blue box), and between BBMs of neurodegeneration-related connectivity and cognitive function in Figure 2B (blue box) (also see Table S3 for numeric data). Regarding AD genetic status, the cluster located in the right Anterior Insula, that showed higher distant connectivity in APOE4 carriers compared with APOE4 non-carriers, was further associated with worse executive function (β = −0.249, p = 0.022), episodic memory (β = −0.229, p = 0.036), processing speed (β = −0.240, p = 0.028) and working memory (β = −0.266, p = 0.014).

Regarding BBMs of neurodegeneration, clusters located in brain regions that showed significant association between NfL levels and higher distant connectivity, were further associated with poorer cognitive function. Specifically, the cluster located in the right Precuneus was associated with worse episodic memory (β = −0.240, p = 0.027), the cluster located in the right Superior Motor Area with worse executive function (β = - 0.287, p = 0.008) and processing speed (β = −0.244, p = 0.024), and the cluster located in the right Putamen with worse processing speed (β = −0.237, 0.029). No significant associations were found for other brain regions or cognitive domains.

## 4. Discussion and Implications

In this cross-sectional study in cognitively normal older adults, we examined how AD pathology, genetic, and BBMs of neurodegeneration relate to patterns of rs dFC, and their implication for cognitive function. Our findings revealed that individuals at higher risk for AD (APOE4 carriers) and those with evidence of pathology (Aβ-positivity), exhibited lower local connectivity in clusters located in frontal regions, while carriers of APOE4 showed higher distant connectivity in clusters located in fronto-insular areas. In relation to BBMs of neurodegeneration, higher levels of NfL, were associated with both, lower local and higher distant connectivity in clusters located in frontal, subcortical and parietal regions. Crucially, several of these distant connectivity patterns were further linked to lower performance in domain-specific cognitive functions. In contrast, we did not observe associations between other BBMs of neurodegeneration such as Aβ40/42 ratios (SIMOA and IPMS), BD-tau, p-tau181, or p-tau217 and rs dFC. Together, these results support the hypothesis that rs dFC disruptions may serve as functional expressions of underlying neurodegenerative processes, reflecting how AD-related biological stages may translate into functional and behavioral alterations.

Rs dFC patterns showed clear distinctions based on AD pathology and genetic status. Concerning local brain connectivity, both Aβ-positive participants and APOE4 carriers presented lower local brain connectivity in the left inferior frontal gyrus compared to their lower-AD risk counterparts. This finding aligns with previous studies suggesting that lower local brain connectivity may reflect disrupted network integrity and less efficient neural processing in individuals with higher neurodegenerative Aβ burden ^36^. Supporting this notion, recent evidence has shown that lower network segregation is associated with higher AD-related tau accumulation among Aβ-positive individuals, highlighting the protective role of maintaining specialized functional organization in early stages of AD ^37^. Given the critical role of frontal regions in executive functioning and working memory ^38^, this pattern may reflect disrupted local communication, potentially undermining cognitive maintenance in the presence of pathological processes.

In contrast to the localized patterns observed for local brain connectivity (associations solely with frontal regions), APOE4 carriers exhibited higher distant brain connectivity in several distributed brain regions, including the left superior motor area, left inferior frontal gyrus, and right anterior insula. This pattern suggests a shift away from localized, specialized processing towards more integrated and less segregated network communication. Although our analysis does not specify the target regions of these distant connections, the increased distal coupling observed may reflect early alterations in large-scale network organization. Previous studies have interpreted higher distant brain connectivity as a potential indicator of dedifferentiation and reduced network specificity, possibly reflecting early inefficiency in functional integration in individuals at greater risk of neurodegeneration ^10,28,33^. Supporting this interpretation, Quevenco et al. (2020) reported that cognitively healthy older adults carrying the APOE4 allele exhibit alterations in functional brain connectivity, particularly in regions vulnerable to early AD pathology, even before the onset of clinical symptoms ^39^. Notably, the superior motor area, inferior frontal gyrus, and anterior insula are part of networks involved in attention, cognitive control, and inter-network regulation ^40^. Altered brain connectivity (i.e., lower local and / or higher distant brain connectivity) in these regions may signal early disruptions in higher-order cognitive functions, consistent with prior evidence linking dysfunctions in executive control, salience, and default mode networks to increased dementia risk and early AD progression ^33,41^.

In addition to pathology and genetic status, we found selective associations for only NfL—among all BBMs of neurodegeneration—showing significant relationships with rs dFC patterns. Specifically, higher levels of NfL were linked to lower local brain connectivity in the right middle cingulate cortex and right insula, as well as higher distant brain connectivity in the right superior motor area, right putamen, right thalamus, left supramarginal gyrus, and right precuneus. This dual pattern (i.e., reduced short-range brain connectivity and increased long-range coupling) may reflect early loss of network segregation and functional specialization, consistent with emerging models of neurodegenerative reorganization. NfL is a well-established marker of axonal injury, and its elevation has been associated with brain atrophy, white matter damage, and disrupted brain connectivity in preclinical stages of AD ^42–44^. Recent longitudinal data further show that individuals with higher levels of NfL exhibit faster declines in functional brain connectivity across several networks, particularly in the presence of amyloid pathology ^45^. Notably, the regions involved in these rs dFC patterns (i.e., cingulate cortex, insula, precuneus, and thalamus) play key roles in attention, memory, and executive control ^40^, suggesting that their disruption may contribute to early inefficiencies in higher-order cognitive processing. The fact that only NfL, and no other BBMs such as Aβ40/42 ratios or phosphorylated tau, showed significant associations with rs dFC might reflect its earlier expression in the neurodegenerative cascade or its closer relationship to axonal integrity and functional network organization ^46,47^.

Beyond the associations between AD risk markers and rs dFC, our results also revealed meaningful links between rs dFC and domain-specific cognitive performance. In particular, APOE4/NfL-related higher distant brain connectivity in regions such as the right anterior insula, right precuneus, right putamen, and right superior motor area were associated with lower scores in executive function, processing speed, episodic memory and working memory. These regions are key nodes within the salience, default mode, and executive control networks, systems critically involved in goal-directed behavior, attentional flexibility, and memory integration. Disruptions in their rs dFC may therefore compromise the brain’s ability to flexibly engage and coordinate functional systems in response to task demands ^48,49^, potentially signaling early network-level mechanisms underlying cognitive decline ^41^. Consistent with these findings, Cao et al. (2021) demonstrated in a longitudinal study that age-related alterations in effective brain connectivity strength within and between key networks, particularly from the left angular gyrus to the precuneus, were predictive of executive function decline in cognitively healthy older adults, underscoring the relevance of network-level dynamics in the aging brain ^50^. Finally, recent evidence from Fischer et al. (2024) indicated that the interplay between AD pathology and genetic status modulates longitudinal functional brain connectivity trajectories and their cognitive correlates, in line with our results that AD risk markers are reflected in both brain connectivity and cognitive performance patterns.

Taken together, our findings revealed a consistent pattern across distinct AD risk markers (i.e., Aβ positivity, APOE4 carriers and higher NfL levels) where increased disease burden was associated with lower local and higher distant rs dFC, and the latter of which was further linked to poorer cognitive function. This shift in rs dFC may signal a breakdown in the brain’s ability to sustain segregated and efficient local processing, accompanied by a compensatory rise in long-range integration aimed at maintaining cognitive function ^8^. While lower local brain connectivity may reflect early alterations in short-range brain connectivity ^33^, the observed higher distant brain connectivity could represent an attempt to recruit broader neural resources ^36,39^. However, such reorganization may come at the cost of network efficiency and specialization ^37^, potentially accelerating cognitive decline over time ^49^. This global pattern supports the hypothesis that rs dFC alterations may serve as sensitive functional markers of early pathological processes ^45^, capturing subtle disruptions in the balance between network segregation and integration that occur before cognitive symptoms become apparent.

This study has some limitations that should be considered. First, the relatively small sample size may limit statistical power and the generalizability of our findings. Second, the cross-sectional design precludes causal interpretations regarding the temporal dynamics between AD risk markers and rs dFC alterations. Longitudinal studies are needed to determine whether the observed brain connectivity associations represent early markers, downstream effects, or compensatory adaptations. Despite these limitations, our study also presents several strengths. Notably, we adopted a whole-brain, data-driven approach to rs dFC, avoiding the constraints of predefined regions or networks and allowing a comprehensive characterization of functional reorganization. Moreover, we integrated a broad panel of AD risk markers, including pathology, genetic, and BBMs of neurodegeneration, providing a multidimensional view of their relationship with rs dFC. Finally, by incorporating domain-specific cognitive measures, we were able to contextualize our findings in terms of cognitive function, enhancing their relevance for early detection and potential intervention strategies.

Our findings provide preliminary evidence that rs dFC may reflect subtle functional alterations associated with early AD–related processes. We observed a consistent brain connectivity pattern characterized by lower local and higher distant brain connectivity across AD risk factors (i.e., Aβ positivity, APOE4 carriers, and higher NfL levels). Interestingly, this pattern of distant brain connectivity was the only one associated with lower cognitive performance, indicating that it could serve as a functional marker of vulnerability to cognitive impairment. These results reinforce the importance of continuing to investigate functional biomarkers that can support earlier detection and risk monitoring. From a public health perspective, better understanding of how rs dFC changes in relation to AD risk may ultimately help guide preventive strategies and promote interventions aimed at preserving cognitive function in aging populations.

## Funding

This work was supported by the Spanish Ministry of Science and Innovation (MCIN/AEI/10.13039/501100011033) and “ERDF A way of making Europe” [grant numbers RTI2018-095284-J-I00, PID2022-137399OB-I00, CNS2024-154835], and by “ESF Investing in your future” [grant number RYC2019-027287-I to I.E-C.]. A.C-P. and M.O-R. are supported by the Spanish Ministry of Science, Innovation and Universities [FPU21/02594 and FPU22/02476, respectively]. This work forms part of the doctoral thesis of A.C-P, conducted within the Biomedicine Doctoral Program at the University of Granada and under the framework of the AGUEDA project.

## Conflict of interests

Kirk I. Erickson has served as a consultant for NeoAuvra, Inc. and MedRhythms, Inc. These affiliations are unrelated to the current study. The remaining authors declare no conflicts of interest.

## Data availability

All scripts, processing pipelines, and study protocols will be made publicly available at the following repository: https://github.com/aguedaprojectugr. The individual-level data used in the present study are not publicly available due to ethical and privacy restrictions, but may be shared upon reasonable request and with appropriate justification.

## Acknowledgements

The authors would like to thank the older adults who participated in this study, the AGUEDA project colleagues, supervisor and co-supervisor for their support. We also thank the co-authors for their valuable contributions and guidance, which improved the quality of this manuscript.

## Authors and coauthors contributions

**Andrea Coca-Pulido:** Writing – original draft, formal analysis, investigation, data curation, **Patricio Solis-Urra:** Project administration, Supervision, Validation, Writing - Review & Editing, **Marcos Olvera-Rojas:** Investigation, data curation, Writing - Review & Editing, **Shivangi Jain:** Writing - Review & Editing, **Anuradha Sehrawat**: Writing - Review & Editing, **Yihun Chen:** Writing - Review & Editing, **Yolanda García-Rivero:** Writing - Review & Editing, **Manuel Gomez-Rio:** Writing - Review & Editing, **Kirk I. Erickson:** Writing - Review & Editing, **Jose Mora-Gonzalez:** Supervision, Writing - Review & Editing, **Irene Esteban-Cornejo**: Funding acquisition, Project administration, Writing - Review & Editing and Supervision.

## Notes

### Clinical Protocols

https://scholar.google.com/citations?view_op=view_citation&hl=es&user=InsxCx4AAAAJ&citation_for_view=InsxCx4AAAAJ:u-x6o8ySG0sC

https://scholar.google.com/citations?view_op=view_citation&hl=es&user=InsxCx4AAAAJ&citation_for_view=InsxCx4AAAAJ:u5HHmVD_uO8C

https://scholar.google.com/citations?view_op=view_citation&hl=es&user=InsxCx4AAAAJ&citation_for_view=InsxCx4AAAAJ:qjMakFHDy7sC

https://scholar.google.com/citations?view_op=view_citation&hl=es&user=InsxCx4AAAAJ&citation_for_view=InsxCx4AAAAJ:zYLM7Y9cAGgC

### Author Declarations

The study followed the Declaration of Helsinki and was approved by the Ethics Board of the Andalusian Health Service (CEIM/CEI Provincial de Granada; #2317 N 19). Written informed consent was obtained from all participants.

## References

1. Livingston G, Huntley J, Liu KY, et al. Dementia prevention, intervention, and care: 2024 report of the Lancet standing Commission. The Lancet.Elsevier B.V. 2024;404(10452):572–628. doi:10.1016/S0140-6736(24)01296-0

2. Jack CR, Andrews JS, Beach TG, et al. Revised criteria for diagnosis and staging of Alzheimer’s disease: Alzheimer’s Association Workgroup. Alzheimers Dement. 2024;20(8):5143–5169. doi:10.1002/alz.13859

3. Yu M, Sporns O, Saykin AJ. The human connectome in Alzheimer disease — relationship to biomarkers and genetics. Nat Rev Neurol.Nature Research. 2021;17(9):545–563. doi:10.1038/s41582-021-00529-1

4. Ashton NJ, Brum WS, Di Molfetta G, et al. Diagnostic Accuracy of a Plasma Phosphorylated Tau 217 Immunoassay for Alzheimer Disease Pathology. JAMA Neurol. 2024;81(3):255–263. doi:10.1001/jamaneurol.2023.5319

5. Wang X, Shi Z, Qiu Y, Sun D, Zhou H. Peripheral GFAP and NfL as early biomarkers for dementia: longitudinal insights from the UK Biobank. BMC Med. 2024;22(1). doi:10.1186/s12916-024-03418-8

6. Yamazaki Y, Zhao N, Caulfield TR, Liu CC, Bu G. Apolipoprotein E and Alzheimer disease: pathobiology and targeting strategies. Nat Rev Neurol. 2019;15(9):501–518. doi:10.1038/s41582-019-0228-7

7. Dennis EL, Thompson PM. Functional brain connectivity using fMRI in aging and Alzheimer’s disease. Neuropsychol Rev. 2014;24(1):49–62. doi:10.1007/s11065-014-9249-6

8. Dai Z, Lin Q, Li T, et al. Disrupted structural and functional brain networks in Alzheimer’s disease. Neurobiol Aging. 2019;75:71–82. doi:10.1016/j.neurobiolaging.2018.11.005

9. Puxeddu MG, Faskowitz J, Betzel RF, Petti M, Astolfi L, Sporns O. The modular organization of brain cortical connectivity across the human lifespan. Neuroimage. 2020;218. doi:10.1016/j.neuroimage.2020.116974

10. Zhao J, Du YH, Ding XT, Wang XH, Men GZ. Alteration of functional connectivity in patients with Alzheimer’s disease revealed by resting-state functional magnetic resonance imaging. Neural Regen Res. 2020;15(2):285. doi:10.4103/1673-5374.265566

11. Ibrahim B, Suppiah S, Ibrahim N, et al. Diagnostic power of resting-state fMRI for detection of network connectivity in Alzheimer’s disease and mild cognitive impairment: A systematic review. Hum Brain Mapp.John Wiley and Sons Inc. 2021;42(9):2941–2968. doi:10.1002/hbm.25369

12. Hansen JY, Shafiei G, Voigt K, et al. Integrating multimodal and multiscale connectivity blueprints of the human cerebral cortex in health and disease. PLoS Biol. 2023;21(9 September). doi:10.1371/journal.pbio.3002314

13. Schulz M, Mayer C, Schlemm E, et al. Association of Age and Structural Brain Changes With Functional Connectivity and Executive Function in a Middle-Aged to Older Population-Based Cohort. Front Aging Neurosci. 2022;14. doi:10.3389/fnagi.2022.782738

14. Tang H, Zhao H, Liu H, et al. Structural damage-driven brain compensation among near-centenarians and centenarians without dementia. Neuroimage. 2025;308. doi:10.1016/j.neuroimage.2025.121065

15. Hartwigsen G. Flexible Redistribution in Cognitive Networks. Trends Cogn Sci.Elsevier Ltd. 2018;22(8):687–698. doi:10.1016/j.tics.2018.05.008

16. Chou Y hui, Chen N kuei, Madden DJ. Functional brain connectivity and cognition: Effects of adult age and task demands. Neurobiol Aging. 2013;34(8):1925–1934. doi:10.1016/j.neurobiolaging.2013.02.012

17. Hausman HK, O’Shea A, Kraft JN, et al. The Role of Resting-State Network Functional Connectivity in Cognitive Aging. Front Aging Neurosci. 2020;12. doi:10.3389/fnagi.2020.00177

18. Chauveau L, Landeau B, Dautricourt S, et al. Anterior-temporal network hyperconnectivity is key to Alzheimer’s disease: from ageing to dementia. Brain. 2025;148(6):2008–2022. doi:10.1093/brain/awaf008

19. Shirer WR, Ryali S, Rykhlevskaia E, Menon V, Greicius MD. Decoding subject-driven cognitive states with whole-brain connectivity patterns. Cerebral Cortex. 2012;22(1):158–165. doi:10.1093/cercor/bhr099

20. Yamashita K ichiro, Uehara T, Prawiroharjo P, et al. Functional connectivity change between posterior cingulate cortex and ventral attention network relates to the impairment of orientation for time in Alzheimer’s disease patients. Brain Imaging Behav. 2019;13(1):154–161. doi:10.1007/s11682-018-9860-x

21. He X, Qin W, Liu Y, et al. Abnormal salience network in normal aging and in amnestic mild cognitive impairment and Alzheimer’s disease. Hum Brain Mapp. 2014;35(7):3446–3464. doi:10.1002/hbm.22414

22. Fernandez-Gamez B, Solis-Urra P, Olvera-Rojas M, et al. Resistance Exercise Program in Cognitively Normal Older Adults: CERT-Based Exercise Protocol of the AGUEDA Randomized Controlled Trial. J Nutr Health Aging. 2023;27(10):885–893. doi:10.1007/s12603-023-1982-1

23. Solis-Urra P, Molina-Hidalgo C, García-Rivero Y, et al. Active Gains in brain Using Exercise During Aging (AGUEDA): protocol for a randomized controlled trial. Front Hum Neurosci. 2023;17. doi:10.3389/fnhum.2023.1168549

24. Sabri O, Sabbagh MN, Seibyl J, et al. Florbetaben PET imaging to detect amyloid beta plaques in Alzheimer’s disease: Phase 3 study. Alzheimer’s and Dementia. 2015;11(8):964–974. doi:10.1016/j.jalz.2015.02.004

25. Barthel H, Gertz HJ, Dresel S, et al. Cerebral amyloid-β PET with florbetaben (18F) in patients with Alzheimer’s disease and healthy controls: a multicentre phase 2 diagnostic study. Lancet Neurol. 2011;10(5):424–435. doi:10.1016/S1474-4422(11)70077-1

26. Sabri O, Seibyl J, Rowe C, Barthel H. Beta-amyloid imaging with florbetaben. Clin Transl Imaging. 2015;3(1):13–26. doi:10.1007/s40336-015-0102-6

27. Minoshima S, Drzezga AE, Barthel H, et al. SNMMI Procedure Standard/EANM Practice Guideline for Amyloid PET Imaging of the Brain 1.0. Journal of Nuclear Medicine. 2016;57(8):1316–1322. doi:10.2967/jnumed.116.174615

28. Diez I, Sepulcre J. Neurogenetic profiles delineate large-scale connectivity dynamics of the human brain. Nat Commun. 2018;9(1). doi:10.1038/s41467-018-06346-3

29. MATLAB Toolbox User’s Guide. The MathWorks Inc. Preprint posted online 2022.

30. Siegel JS, Power JD, Dubis JW, et al. Statistical improvements in functional magnetic resonance imaging analyses produced by censoring high□motion data points. Hum Brain Mapp. 2014;35(5):1981–1996. doi:10.1002/hbm.22307

31. Oberlin LE, Wan L, Kang C, et al. Cardiorespiratory fitness is associated with cognitive function in late adulthood: baseline findings from the IGNITE study. Br J Sports Med. Published online December 10, 2024:bjsports-2024-108257. doi:10.1136/bjsports-2024-108257

32. Dong Y, Peng CYJ. Principled missing data methods for researchers. Springerplus. 2013;2(1):222. doi:10.1186/2193-1801-2-222

33. Dautricourt S, Gonneaud J, Landeau B, et al. Dynamic functional connectivity patterns associated with dementia risk. Alzheimers Res Ther. 2022;14(1). doi:10.1186/s13195-022-01006-7

34. Viviano RP, Raz N, Yuan P, Damoiseaux JS. Associations between dynamic functional connectivity and age, metabolic risk, and cognitive performance. Neurobiol Aging. 2017;59:135–143. doi:10.1016/j.neurobiolaging.2017.08.003

35. Roemer-Cassiano SN, Wagner F, Evangelista L, et al. Amyloid-associated hyperconnectivity drives tau spread across connected brain regions in Alzheimer’s disease. Sci Transl Med. 2025;17(782). doi:10.1126/scitranslmed.adp2564

36. Franzmeier N, Neitzel J, Rubinski A, et al. Functional brain architecture is associated with the rate of tau accumulation in Alzheimer’s disease. Nat Commun. 2020;11(1):347. doi:10.1038/s41467-019-14159-1

37. Steward A, Biel D, Brendel M, et al. Functional network segregation is associated with attenuated tau spreading in Alzheimer’s disease. Alzheimer’s & Dementia. 2023;19(5):2034–2046. doi:10.1002/alz.12867

38. Stuss DT. Functions of the frontal lobes: Relation to executive functions. Journal of the International Neuropsychological Society.Cambridge University Press. 2011;17(5):759–765. doi:10.1017/S1355617711000695

39. Quevenco FC, van Bergen JM, Treyer V, et al. Functional Brain Network Connectivity Patterns Associated With Normal Cognition at Old-Age, Local β-amyloid, Tau, and APOE4. Front Aging Neurosci. 2020;12. doi:10.3389/fnagi.2020.00046

40. Ji JL, Spronk M, Kulkarni K, Repovš G, Anticevic A, Cole MW. Mapping the human brain’s cortical-subcortical functional network organization. Neuroimage. 2019;185:35–57. doi:10.1016/j.neuroimage.2018.10.006

41. Badhwar AP, Tam A, Dansereau C, Orban P, Hoffstaedter F, Bellec P. Resting-state network dysfunction in Alzheimer’s disease: A systematic review and meta-analysis. *Alzheimer’s and Dementia: Diagnosis*, Assessment and Disease Monitoring. 2017;8:73–85. doi:10.1016/j.dadm.2017.03.007

42. Jung Y, Damoiseaux JS. The potential of blood neurofilament light as a marker of neurodegeneration for Alzheimer’s disease. Brain. 2024;147(1):12–25. doi:10.1093/brain/awad267

43. Khalil M, Pirpamer L, Hofer E, et al. Serum neurofilament light levels in normal aging and their association with morphologic brain changes. Nat Commun. 2020;11(1):812. doi:10.1038/s41467-020-14612-6

44. Brier MR, Day GS. Serum neurofilament light chain uncovers neurodegeneration early in the course of Alzheimer’s disease. Brain. 2020;143(12):3521–3522. doi:10.1093/brain/awaa370

45. Dark HE, Shafer AT, Cordon J, et al. Association of Plasma Biomarkers of Alzheimer Disease and Neurodegeneration With Longitudinal Intra-Network Functional Brain Connectivity. Neurology. 2025;104(4). doi:10.1212/WNL.0000000000210271

46. Leuzy A, Mattsson□Carlgren N, Palmqvist S, Janelidze S, Dage JL, Hansson O. Blood□based biomarkers for Alzheimer’s disease. EMBO Mol Med. 2022;14(1). doi:10.15252/emmm.202114408

47. Hansson O, Blennow K, Zetterberg H, Dage J. Blood biomarkers for Alzheimer’s disease in clinical practice and trials. Nat Aging.Springer. 2023;3(5):506–519. doi:10.1038/s43587-023-00403-3

48. Quevenco FC, Preti MG, van Bergen JMG, et al. Memory performance-related dynamic brain connectivity indicates pathological burden and genetic risk for Alzheimer’s disease. Alzheimers Res Ther. 2017;9(1):24. doi:10.1186/s13195-017-0249-7

49. Staffaroni AM, Brown JA, Casaletto KB, et al. The longitudinal trajectory of default mode network connectivity in healthy older adults varies as a function of age and is associated with changes in episodic memory and processing speed. Journal of Neuroscience. 2018;38(11):2809–2817. doi:10.1523/JNEUROSCI.3067-17.2018

50. Cao X, Liu T, Jiang J, et al. Alternation in Effective Connectivity With Cognitive Aging: A Longitudinal Study of Elderly Populations. Front Aging Neurosci. 2021;13. doi:10.3389/fnagi.2021.755931

