## Supplementary Tables 1-3 for "Alzheimer’s risk markers and resting-state dynamic functional connectivity: Cross-Sectional Findings from the AGUEDA Study"

**Supplementary Materials**

| **Supplementary Table 1.** Differences in resting-state dynamic functional connectivity among Alzheimer Disease pathology / genetic status in cognitively normal older adults (n = 86) | | | | | | | | | | | | | | | | | | | |
| --- | --- | --- | --- | --- | --- | --- | --- | --- | --- | --- | --- | --- | --- | --- | --- | --- | --- | --- | --- |
| **Alzheimer Disease pathology/genetic status variables** | **Local Connectivity** | | | | | | | | |  | **Distant Connectivity** | | | | | | | | |
|  | **Brain region** | **x** | **y** | **z** | **k** | **t** | **β** | **p(uncorrected)** | **pFWE corrected** |  | **Brain region** | **x** | **y** | **z** | **k** | **t** | **β** | **p(uncorrected)** | **pFWE corrected** |
| **APOE4 (carriers vs non-carriers)** |  | | | | | | | | | | | | | | | | | | |
| *non-carriers>carriers* | Inferior Frontal Gyrus | -52 | 10 | 22 | 40 | -4.05 | -0.215 | 0.000 | 0.003 |  | - | | | | | | | | |
| *carriers>non-carriers* | - | | | | | | | | |  | Superior Motor Area | -4 | 4 | 58 | 73 | 3.83 | 0.577 | 0.000 | 0.000 |
|  | - | | | | | | | | |  | Inferior Frontal Gyrus | -52 | 10 | 16 | 102 | 4.62 | 0.65 | 0.000 | 0.000 |
|  | - | | | | | | | | |  | Anterior Insula | 38 | 28 | 4 | 41 | 5.62 | 0.682 | 0.000 | 0.011 |
| **Aβ status (>=12 centiloid)** |  | | | | | | | | | | | | | | | | |  |  |
| *negative>positive* | Inferior Frontal Gyrus | -52 | 10 | 22 | 56 | -4.07 | -0.047 | 0.000 | 0.001 |  | - | | | | | | | | |
| *positive>negative* | - | | | | | | | | |  | - | | | | | | | | |
| Note. Local and distal dynamic functional connectivity maps obtained for each individual were entered in regression linear models to determine group differences across AD pathology / genetic status (predictor variables) controlling for age, sex and education. Five participants were defined as outliers and excluded from the analyses due to having more than 25% of invalid volumes in the fMRI sequence. Statistical significance was set at a peak-level p-value (uncorrected) ≤ 0.01 and a pFWE(corrected) cluster level < 0.05 to assess differences between groups. Anatomical coordinates (X, Y, Z) are given in Montreal Neurological Institute (MNI) Atlas space. K value represents the cluster size. T values reflect the magnitude and direction of group differences. β values represents the standardized beta coefficients. APOE4 allele status: carriers vs non-carriers. Aβ status: amyloid-β positive vs negative. | | | | | | | | | | | | | | | | | | | |

| **Supplementary Table 2.** Association between blood-based markers of neurodegeneration and clusters located in brain regions with stable local and distant connectivity patterns (attractors) through resting state dynamic functional connectivity in cognitively normal older adults (n = 86) | | | | | | | | | | | | | | | | | | | |
| --- | --- | --- | --- | --- | --- | --- | --- | --- | --- | --- | --- | --- | --- | --- | --- | --- | --- | --- | --- |
| **Blood-based markers of neurodegeneration** | **Local Connectivity** | | | | | | | | |  | **Distant Connectivity** | | | | | | | | |
|  | **Brain region** | **x** | **y** | **z** | **k** | **t** | **β** | **p(uncorrected)** | **pFWE corrected** |  | **Brain region** | **x** | **y** | **z** | **k** | **t** | **β** | **p(uncorrected)** | **pFWE corrected** |
| **Aβ40/42 ratio simoa** | - | | | | | | | | |  | - | | | | | | | | |
| **Aβ40/42 ratio ipms** | - | | | | | | | | |  | - | | | | | | | | |
| **Brain-derived tau** | - | | | | | | | | |  | - | | | | | | | | |
| **GFAP pgml simoa** | - | | | | | | | | |  | - | | | | | | | | |
| **NfL pgml simoa** | - | | | | | | | | |  | - | | | | | | | | |
| *Positive Contrast* | - | | | | | | | | |  | Superior Motor Area | 8 | -2 | 46 | 98 | 3.69 | 0.389 | 0.000 | 0.000 |
|  | - | | | | | | | | |  | Putamen | 32 | -2 | 4 | 181 | 4.16 | 0.439 | 0.000 | 0.000 |
|  | - | | | | | | | | |  | Thalamus | 2 | -8 | 10 | 30 | 3.89 | 0.423 | 0.000 | 0.048 |
|  | - | | | | | | | | |  | Supramarginal | -64 | -38 | 28 | 119 | 3.67 | 0.402 | 0.000 | 0.000 |
|  | - | | | | | | | | |  | Precuneus | 2 | -56 | 52 | 40 | 4.11 | 0.43 | 0.000 | 0.012 |
| *Negative Contrast* | Middle Cingulate | 2 | -8 | 40 | 28 | -3.8 | -0.414 | 0.000 | 0.027 |  | - | | | | | | | | |
|  | Insula | 38 | -14 | 16 | 38 | -4.27 | -0.453 | 0.000 | 0.005 |  | - | | | | | | | | |
| **P-tau181** | - | | | | | | | | |  | - | | | | | | | | |
| **P-tau217** | - | | | | | | | | |  | - | | | | | | | | |
| Note. Local and distal resting state dynamic functional connectivity maps obtained for each individual were entered in regression linear models to determine the association with blood-based markers of neurodegeneration (predictor variables) controlling for age, sex and education. Five participants were defined as outliers and excluded from the analyses due to having more than 25% of invalid volumes in the fMRI sequence. Statistical significance was set at a peak-level p-value (uncorrected) ≤ 0.01 and a pFWE(corrected) cluster level <0.05 to assess the association between blood-based markers of neurodegeneration and connectivity maps. Anatomical coordinates (X, Y, Z) are given in Montreal Neurological Institute (MNI) Atlas space. All markers are interpreted as the lower values, the better. Data for one participant was not available for the variables Aβ40/42 ratio, NfL, and GFAP, while data for another participant was missing for the variable p-tau217. As a result, each analysis was conducted with 85 participants.  Abbrebiations: Aβ40/42 ratio: β-amyloid 1-42 to β-amyloid 1-40 ratio. P-tau217 (pg/ml): Concentration of phosphorylated tau protein at position 217 in the brain, measured in picograms per milliliter. P-tau181 (pg/ml): Concentration of phosphorylated tau protein at position 181. NfL (pg/ml) SIMOA: Levels of neurofilament light chain (NFL). GFAP (pg/ml) SIMOA: Concentration of glial fibrillary acidic protein (GFAP). | | | | | | | | | | | | | | | | | | | |

| **Supplementary Table 3.** Alzheimer disease (AD) pathology / genetic status and blood-based markers of neurodegeneration-related connectivity attractorness and their association with cognitive function in cognitively normal older adults (n = 86) | | | | | | | |
| --- | --- | --- | --- | --- | --- | --- | --- |
| AD risk markers | Brain region (Connectivity) | Executive Function | Episodic Memory | Processing Speed | Working Memory | Attentional Control | Visuospatial Processing |
|  |  | *β* | *β* | *β* | *β* | *β* | *β* |
| APOE4 | Inferior frontal gyrus (Lc) | 0.022 | -0.007 | -0.083 | 0.046 | -0.009 | -0.025 |
|  | Frontal Inferior Operculum (Dc) | -0.05 | -0.148 | -0.027 | -0.042 | 0.008 | 0.022 |
|  | Anterior Insula (Dc) | **-0.249*** | **-0.229*** | **-0.240*** | **-0.266*** | -0.072 | -0.168 |
|  | Superior Motor Area (Dc) | -0.022 | -0.172 | -0.010 | -0.05 | 0.05 | 0.042 |
| Aβ status | Inferior frontal gyrus (Lc) | 0.029 | -0.009 | -0.077 | 0.054 | 0.005 | -0.031 |
| NfL | Middle Cingulate (Lc) | 0.032 | 0.037 | 0.046 | -0.012 | -0.043 | -0.069 |
|  | Insula (Lc) | -0.085 | 0.091 | -0.080 | -0.089 | -0.095 | -0.043 |
|  | Precuneus (Dc) | -0.073 | **-0.240*** | -0.140 | -0.006 | 0.001 | 0.105 |
|  | Putamen (Dc) | -0.189 | -0.195 | **-0.237*** | -0.201 | -0.118 | -0.109 |
|  | Superior Motor Area (Dc) | **-0.287*** | -0.127 | **-0.244*** | -0.172 | -0.149 | 0.019 |
|  | Supramarginal (Dc) | -0.126 | -0.050 | -0.094 | -0.092 | -0.043 | 0.022 |
|  | Thalamus (Dc) | -0.018 | -0.016 | -0.083 | 0.036 | 0.099 | 0.195 |
| Note. Values are standardized regression coefficients (β). Statistically significant values (marked in **bold***) were set at (P ≤ 0.05). Data for one participant was not available for Executive Function. As a result, the analysis was conducted with one less participant. Executive function and cognitive subdomains are expressed as z-scores.  Abbrebiations: AD: Alzheimer Disease; APOE4: carriers / non-carriers; Aβ status: positive / negative; Dc: Distant Connectivity; NfL: Neurofilament Light chain. Lc: Local Connectivity.  * p < .05. | | | | | | | |
